# SEXUAL MISCONDUCT AMONG HIGH SCHOOL STUDENTS IN VIETNAM

**DOI:** 10.1101/2025.01.28.25321240

**Authors:** Kim Tu Tran, Ruschelle M. Leone, Kevin M Swartout, Minh Hung Tran, Oanh Trinh, Kathryn M. Yount

## Abstract

**Background:** Sexual misconduct is a global problem. Adolescents 15-19 years face the highest risk of sexual misconduct, however, studies on school-based sexual misconduct in low and middle-income countries (LMICs) are limited. We aimed to determine the prevalence of sexual misconduct, including sexual harassment, stalking, dating violence, and sexual violence experiences among students at three high schools in Vietnam since their enrollment.

**Methods:** Between February and May 2023, 754 students in three high schools in Ho Chi Minh City completed the adapted Administrator-Researcher Campus Climate Collaborative online.

**Results:** Overall, 54.5% of students reported any sexual misconduct since enrolling in high school. The prevalence of sexual harassment victimization was 40.2% perpetrated by staff and 30.2% perpetrated by students. Stalking prevalence was 18.3%, and 13.1% of students reported experiencing dating violence. Nearly 1 in 10 (8.7%) of students reported sexual violence victimization. Sexual coercion prevalence was highest in the middle-ranked school (6.2%) and lowest in the high-ranked school (1.6%) (p=0.028). Compared to girls, boys reported a higher prevalence of unwanted sexual attention (18.7% vs. 10.9%) and sexual coercion (10.1% vs 4.6%) by staff (p=0.001). Girls reported a higher prevalence of dating violence victimization than boys (14.9% vs. 9.9%, p=0.029).

**Conclusion:** Sexual misconduct prevalence among high school students in Ho Chi Minh City is high with notable gender differences in victimization. Awareness raising among parents, school administrators, and youth is needed to develop contextually appropriate preventive and response programs to address sexual misconduct in high schools across Vietnam.

**Highlights:**

- We assessed sexual misconduct among students in three high schools in South Vietnam
- We assessed experiences of sexual harassment, stalking, dating violence, and sexual violence
- Boys experienced higher sexual harassment victimization by faculty/staff than girls
- Girls experienced a higher prevalence of dating violence victimization than boys
- Findings inform future research and prevention efforts for high schools in South Vietnam

## INTRODUCTION

Sexual misconduct is a global problem. Sexual misconduct refers to “physical or non-physical conduct of a sexual nature, including sexual harassment, stalking, dating violence, and sexual violence in the absence of clear, knowing, affirmative consent” (Swartout et al., 2019) (p. 9). Studies have shown that adolescents 15-19 years of age are at the highest risk for sexual misconduct (Sahu et al., 2005), and adolescent girls in this age group are four times more likely to experience sexual misconduct than girls in other age groups (Banvard-Fox et al., 2020). The latest report of the Youth Risk Behavior Survey in 2022 showed that students in U.S. high schools, grades 9 through 12, experienced a higher rate of sexual misconduct than secondary school students, specifically those in grades 6 and 8 (Emerson Hospital, 2022). Further, among students, girls are six times more likely than boys to be victims of sexual misconduct (Emerson Hospital, 2022).

Few studies have investigated sexual misconduct in high schools, with limited data on different forms of sexual misconduct. A nationally representative study of 2,200 students in grade 10 in the U.S. showed 40% of girls and 27% of boys experienced dating violence victimization (Haynie et al., 2013). In one of the first population-based studies of 18,000 high-school students in the U.S., a higher percentage of girls than boys experienced sexual misconduct in the past 12 months, for example, being stalked (19% vs. 14%) (Fisher et al., 2014), being sexually harassed (37% vs. 21%) (Clear et al., 2014), and being exposed to sexual violence (23% vs. 14%) (Williams et al., 2014).

In Asia, few studies have investigated adolescents’ exposure to sexual misconduct in schools. In a 2017 study of 3,500 middle-school and high-school students in Indonesia, 4.5% reported sexual violence (Syukriani et al., 2022); however, the items asked in the questionnaire were unclear, precluding interpretation of this prevalence rate. In China, a national survey of 9,000 secondary-school students, grades 7 through 12, found that 13% reported at least one experience of sexual violence in childhood, with reporting highest for sexual harassment (6.0%) and sexual violence (6.9%) (Zhang et al., 2021). In Vietnam, a study conducted in 2014 among 2,300 students 12-17 years old from secondary and high schools found that 20% reported experiencing sexual violence (Tran et al., 2018). However, only eight questions about sexual violence were asked, which may not capture the full range of experiences of contact and non-contact forms of sexual violence among students. Similar to the study in China, the study in Vietnam asked about lifetime sexual misconduct, which is not specific to school-based exposure to sexual misconduct.

Adolescence is a critical period of development, and exposure to sexual misconduct during adolescence has a range of adverse effects on physical, psychological, social, and health outcomes and behaviors. Victims of child abuse are at heightened risk of revictimization in intimate relationships later in life (Schuster & Tomaszewska, 2021). Mental health effects including depression, suicidal ideation, and attempted suicide are frequently reported in victims of sexual misconduct (Brokke et al., 2022). Victims of sexual misconduct are more likely to exhibit unhealthy coping behaviors, such as alcohol binge and drug abuse (Rich et al., 2016), which may themselves contribute to risky sexual behavior (Solehati et al., 2021).

Given the adverse effects of sexual misconduct on victims and limited data on experiences of sexual misconduct among high-school students in Vietnam, this pilot study provides the first comprehensive estimates for the prevalence rates of specific forms of sexual misconduct among high-school students in Vietnam. This pilot study also provides insights about the feasibility and acceptability of conducting high-school based interventions to prevent the most prevalent forms of violence against high-school students and will inform the development or adaptation of specific high-school violence prevention interventions that address these forms of violence.

## METHODS

### Participants

The survey was conducted in May 2023 at three high schools in Ho Chi Minh city. One school each was selected randomly from within ‘good’ rank, ‘middle’ rank, and ‘low’ rank schools, based on the national entrance examination test scores. High school in Vietnam includes grade 10 (15 years old), grade 11 (16 years old), and grade 12 (17 years old). To capture experiences that occurred in high school, we recruited students in grades 11 and 12, as these students had been enrolled in high school for at least 12 months. Grade 10 students were excluded because they had yet to be in high school long enough to report on school-based experiences of sexual misconduct in the prior 12 months.

We estimated a sample size of 246 students per school, based on a 20% estimated prevalence rate of sexual misconduct (Tran et al., 2018) and a relative precision of 5%. Based on a prior study (Underwood et al., 2020), we assumed that item-level missingness would be 40% (in other words, 40% of participants would not answer questions on sexual misconduct). Therefore, we aimed to recruit 345 students per school. Students in each grade were selected based on probability proportional to the total number of students in that grade, and classes covering the necessary number of students were selected randomly.

### Data

The questionnaire was adapted and expanded from the Administrative Researcher Campus Climate Collaborative (ARC3) survey (Swartout et al., 2019), a free campus climate survey designed by sexual assault researchers and administrators for U.S. institutions of higher education. The original ARC3 survey was designed to assess demographic characteristics, alcohol use, and forms of sexual misconduct identified as Title IX violations, including sexual harassment, stalking, dating violence, and sexual violence victimization and perpetration since students enrolled in school. Each domain of sexual misconduct in the ARC3 has exhibited good psychometric properties among students attending seven universities in the U.S. (Tilley et al., 2020).

Experts on sexual violence in Vietnam reviewed the ARC3 survey and recommended modules that were likely to be culturally suitable for high school students in this setting. The study team also considered the time required for adolescents to self-administer an adapted online survey to support questionnaire completion. The final adapted survey included six modules. A *demographics module* combined questions on sex at birth, age, gender, ethnicity, sexual orientation, grade, organizational members, and living arrangements from modules in the ARC3 survey (Swartout et al., 2019) and a self-administered online survey with undergraduate men in Vietnam (Yount et al., 2023). Four selected ARC3 modules measured sexual harassment victimization, stalking victimization, dating violence victimization, and sexual violence victimization. The final module, originally from the GlobalConsent efficacy trial (Yount et al., 2023), asked about the frequency of using sexually explicit media and sexually violent media, as sexually violent media use is associated with sexually violent behavior among undergraduates in Vietnam (Bergenfeld, Cheong, et al., 2022). More detail on the selected ARC3 modules is warranted, given the focus of this analysis.

The module on *sexual harassment victimization by faculty or staff* includes four sections and 16 items originally from the Sexual Experiences Questionnaire (Fitzgerald et al., 1999) measuring the frequency of sexist hostility, sexual hostility, unwanted sexual attention, and sexual coercion. In this sample, internal consistency was 0.88-0.95 for items in each sub-construct and 0.95 for all items in this set. The module on *sexual harassment victimization by students,* also originally from the Sexual Experiences Questionnaire, includes four sections with 16 items measuring the frequency of sexist hostility, sexual hostility, unwanted sexual attention, and sexual harassment via e-communications conducted by students. Internal consistency for each subset of items was 0.87-0.92 and for the full set of items was 0.95 in this sample. The module on *stalking victimization*, originally from the National Intimate Partner and Sexual Violence Survey (NISVS) (Centers for Disease Control and Prevention, 2011), includes 10 items assessing how often students have been purposefully pursued by someone since enrolling at school. Internal consistency for this item set was 0.92 in this sample. The module on *dating violence victimization*, originally from the Partner Victimization Scale (Hamby, 2014) and the Women’s Experience with Battering Scale (Smith et al., 1995), includes six items assessing the frequency of physical and psychological dating violence conducted by a current or former boyfriend or girlfriend since enrollment at school. Internal consistency for this item set was 0.92 in this sample. The module on *sexual violence*, originally from the Sexual Experiences Survey (Koss et al., 1987), includes four sections and 25 items that measure the frequency of: (1) sexual contact (e.g., nonconsensual fondling, kissing, rubbing private areas, or removing clothes), (2) attempted sex (e.g., attempted coercion or rape though not successful, (3) coercion (being forced into oral sex or intercourse through verbal threats), and (4) rape (being forced into oral sex or intercourse through physical threats). Internal consistency was 0.74-0.96 across item subsets in this sample. All ARC3 questions on sexual misconduct were asked with respect to the period since the students’ enrollment in high schools.

### Procedures

Parents and students of selected classes were provided consent and assent by research assistants to participate. Students who assented to participate and had a parent consent to their participation were given the survey questionnaire. A voucher of $3 was sent promptly to students upon completing the survey as a token of appreciation.

The survey was delivered to students via a secure link using the REDCap data collection system. REDCap is a data system that allows for the secure (encrypted) entry, transfer, and storage of data and user-defined access to data elements. The survey was anonymous, and no identifiers were collected to protect the confidentiality of students and to ensure honest responses. Students accessed the survey link using their mobile phones with 3G connectivity. Two non-study study staff were present in class to assist participants with any technological issues with the survey, as needed. The survey link address was altered after each data collection to prevent students from re-accessing it and adding additional records. The study received ethical approval from the Institutional Review Board of the University of Medicine and Pharmacy at Ho Chi Minh city.

### Outcomes

The outcomes for this analysis focused on experiences of sexual misconduct only, given the paucity of research on this topic among high-school students in Vietnam. Sexual harassment was recoded 1 if a participant reported to have experienced any form of sexual harassment since enrolling in high school, 0 if the participant reported none, and left blank if the participant did not respond to any items on sexual harassment. Stalking victimization was recoded 1 if a participant reported to have experienced any form of stalking victimization since enrolling in high school, 0 if none were reported, and left blank if responses to all stalking items were missing. Dating violence victimization was recoded 1 if the participant reported to have experienced any form of dating violence since enrolling in high school, 0 if no form was reported, and left blank if all items on dating violence were missing. Sexual violence victimization was recoded 1 if students reported to have experienced any item on unwanted sexual contact, attempted sex, coercion, or rape since enrolling in school, 0 if the participant reported none, and left blank if they did not respond to any item on sexual violence victimization.

### Covariates

The primary covariates for comparing sexual misconduct prevalence rates were school (high, medium, low rank) and cis-gender self-identification (self-identified male, self-identified female). Very few participants reported a non-cis-gender identification or did not report their gender (N≈34), and those participants were dropped from the main outcomes-related analysis.

### Analysis

This descriptive analysis followed three primary steps. First, given the pilot nature of this study, overall participation rates were computed for the pooled sample to understand the overall feasibility and acceptability of conducting a climate survey of this nature and scope with high-school students in Vietnam. The demographic characteristics of the sample then were assessed overall and by school and gender to understand how these subsamples may have differed on characteristics that also may be associated with experiences of sexual misconduct. Univariate analysis then was conducted with all original sexual misconduct items to understand their distributions and level of missingness. We then estimated the prevalence rates since enrolling in school, for the overall sample, by school, and by gender for reported experiences of: sexual harassment by faculty/staff; sexual harassment by students; stalking; dating violence, and sexual violence. Stata 14 version was used to process and to analyze all data.

## RESULTS

### Participation in the climate survey

Out of 1012 students eligible for the study, 754 participated, resulting in an overall participation rate of 74.5%. Participation rates varied modestly across schools (78% to 84%) and across gender (82% by girls versus 79% by boys). Notably, the total number of received survey records was 934. Due to internet disruptions, some students lost access to their survey and created another record, resulting in the final number of observations in the dataset exceeding the actual number of participants by 180 (19.3%) due to duplicate records. Duplicate records could not be removed because no identifiers were collected. Therefore, the analysis was performed on 934 records. The mean time to complete the survey is 20:23.

### Participant demographics

About one third of the sample came from each school (33.8%, 33.4%, and 32.8%) (Table 1). The percentage of participants in grade 12 was higher than that in grade 11 (55.1% vs. 44.9%). Most participants were Vietnamese (96.4%). A slight majority of participants self-identified as cis-female (56.2%), and the remainder self-identified as cis-male (40.2%), non-binary (2.3%), or transgender (0.6%), with 0.7% not responding to the question on gender. Most participants identified as heterosexual (74.6%), although a sizable minority self-identified as a sexual minority. Most participants reported living with their parents (90.3%).

**Table 1.**
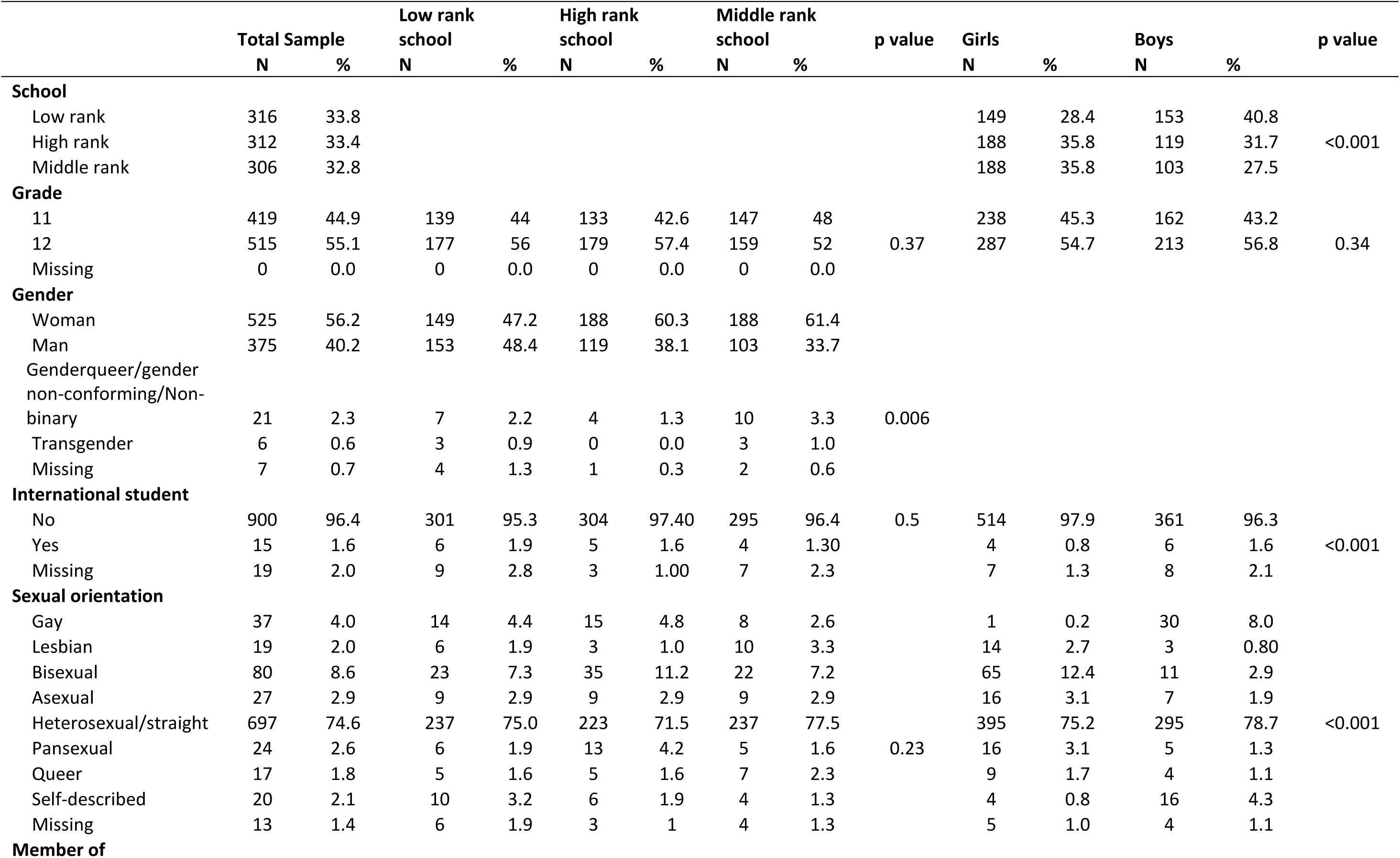
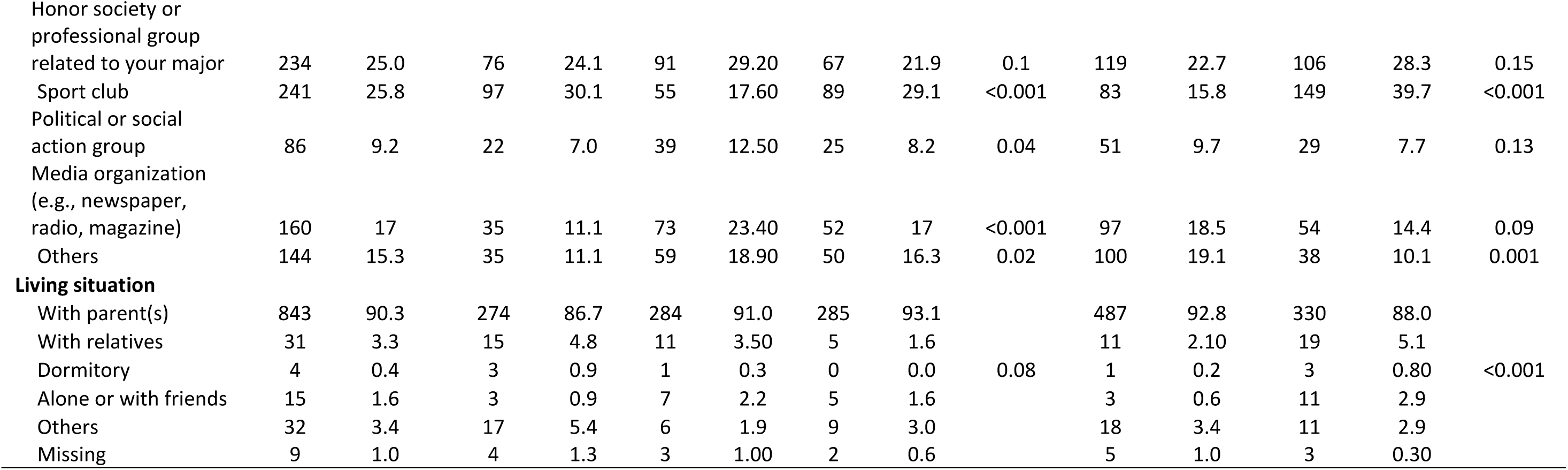
Demographic Characteristics of the Sample, students in grades 11 and 12 in three high schools in Ho Chi Minh City.

### Reports of sexual harassment victimization

The prevalence rate for sexual harassment victimization by faculty or staff was 40.2%, with the sub-domain of sexist hostility being most prevalent (30.2%), followed by sexual hostility (27.1%), unwanted sexual attention (15.0%), and sexual coercion (7.6%) (Table 2).

**Table 2:** Sexual misconduct among students in grades 11 and 12 in three high schools in Ho Chi Minh City.

|  | N | n | Prevalence | Missing | alpha |
| --- | --- | --- | --- | --- | --- |
| **Sexual Harassment Victimization by faculty/staff** | 805 | 376 | 40.2 | 13.8 | 0.95 |
| Sexist hostility | 836 | 282 | 30.2 | 10.5 | 0.88 |
| Sexual hostility | 835 | 253 | 27.1 | 10.6 | 0.88 |
| Unwanted sexual attention | 835 | 140 | 15.0 | 10.6 | 0.91 |
| Sexual coercion | 823 | 71 | 7.6 | 11.9 | 0.95 |
| **Sexual Harassment Victimization by students** | 783 | 320 | 34.3 | 16.2 | 0.95 |
| Sexist hostility | 817 | 180 | 19.3 | 12.5 | 0.90 |
| Sexual hostility | 811 | 225 | 24.1 | 13.2 | 0.91 |
| Unwanted sexual attention | 809 | 171 | 18.3 | 13.4 | 0.92 |
| Sexual harassment via e-communication | 804 | 174 | 18.6 | 13.9 | 0.87 |
| **Stalking Victimization** | 791 | 171 | 18.3 | 15.3 | 0.92 |
| **Dating Violence Victimization** | 737 | 122 | 13.1 | 21.1 | 0.92 |
| **Sexual violence victimization** |  |  |  |  |  |
| Sexual contact | 783 | 60 | 6.4 | 16.2 | 0.86 |
| Attempted sex | 776 | 26 | 2.8 | 16.9 | 0.92 |
| Attempted coercion | 782 | 20 | 2.1 | 16.3 | 0.74 |
| Attempted rape | 777 | 20 | 2.1 | 16.8 | 0.90 |
| Coercion | 735 | 38 | 4.1 | 21.3 | 0.91 |
| Rape | 722 | 30 | 3.2 | 22.7 | 0.96 |
| Any sexual violence victimization | 794 | 81 | 8.7 | 15.0 |  |
| **Any sexual misconduct** | 748 | 509 | 54.5 | 19.9 |  |

The overall prevalence of sexual harassment victimization by faculty or staff did not differ by school or by gender (Table 3). However, the prevalence rates for two sub-domains of sexual harassment— namely unwanted sexual attention (18.7% and 10.9%) and sexual coercion (10.1% and 4.6%)—were significantly higher for boys than girls, respectively (Table 4).

**Table 3:**
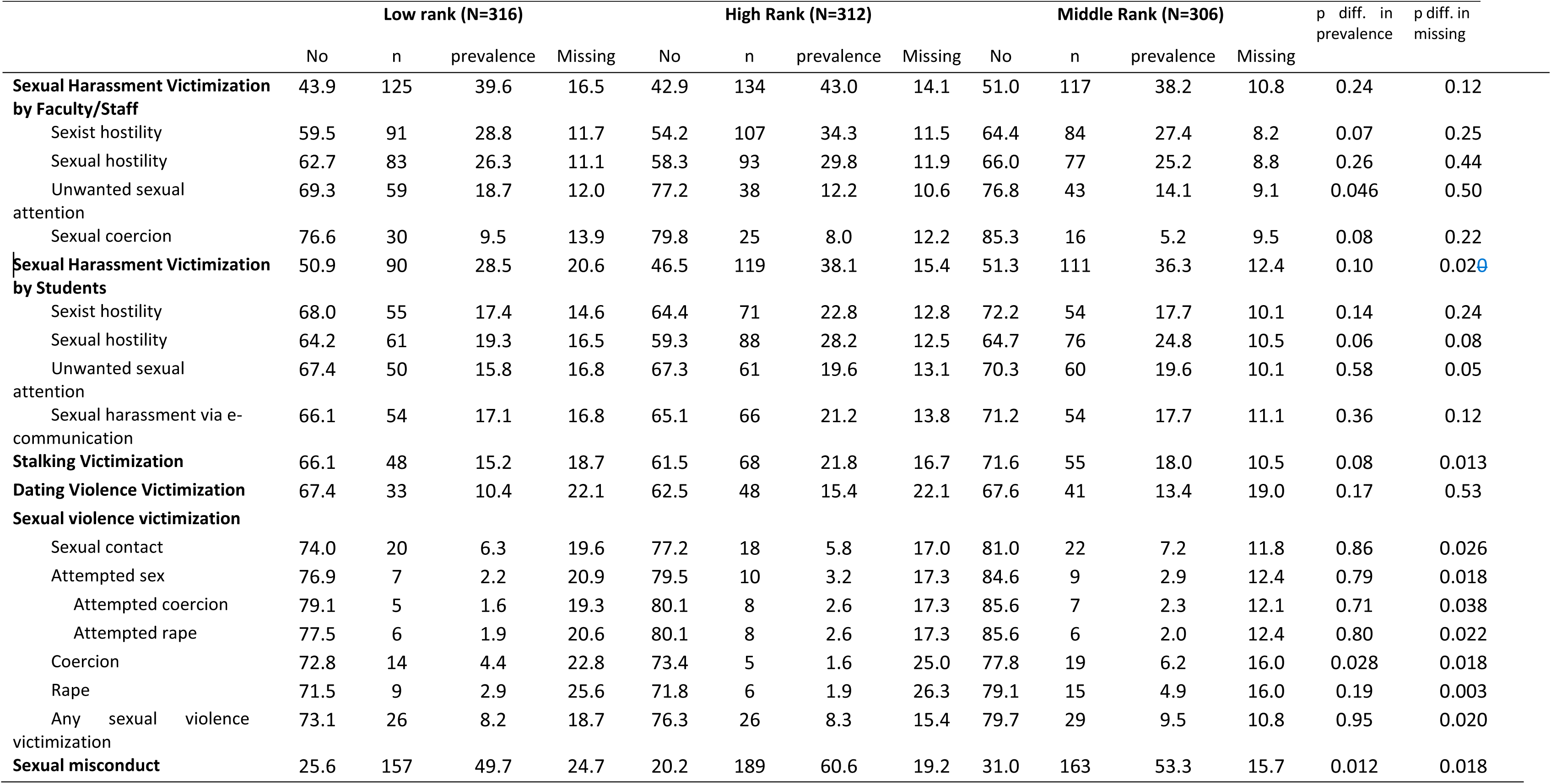
Sexual misconduct among students in grades 11 and 12 in three high schools in Ho Chi Minh City, by type of school.

**Table 4:** Sexual misconduct among students in grades 11 and 12 in three high schools in Ho Chi Minh City, by gender.

|  | Girls<br>(N=525) |  |  |  | Boys<br>(N=375) |  |  |  | p diff. in<br>prevalence | p diff. In<br>missing |
| --- | --- | --- | --- | --- | --- | --- | --- | --- | --- | --- |
|  | No | n | prevalence | Missing | No | n | prevalence | Missing |  |  |
| **Sexual Harassment Victimization by faculty/staff** | 47.2 | 205 | 39.1 | 13.7 | 45.8 | 153 | 40.8 | 13.3 | 0.61 | 0.86 |
| Sexist hostility | 61.5 | 148 | 28.2 | 10.3 | 57.8 | 118 | 31.50 | 10.7 | 0.25 | 0.85 |
| Sexual hostility | 63.0 | 141 | 26.9 | 10.1 | 62.9 | 98 | 26.10 | 10.9 | 0.87 | 0.68 |
| Unwanted sexual attention | 79.2 | 57 | 10.9 | 9.9 | 69.9 | 70 | 18.7 | 11.5 | 0.001 | 0.73 |
| Sexual coercion | 84.0 | 24 | 4.6 | 11.4 | 77.9 | 38 | 10.1 | 12.0 | 0.001 | 0.55 |
| **Sexual Harassment Victimization by students** | 48.6 | 182 | 34.7 | 16.8 | 51.7 | 123 | 32.8 | 15.5 | 0.43 | 0.60 |
| Sexist hostility | 69.3 | 97 | 18.5 | 12.2 | 67.5 | 73 | 19.5 | 13.1 | 0.65 | 0.91 |
| Sexual hostility | 63.8 | 123 | 23.4 | 12.8 | 62.1 | 91 | 24.3 | 13.6 | 0.70 | 0.71 |
| Unwanted sexual attention | 69.10 | 96 | 18.3 | 12.6 | 68.0 | 66 | 17.6 | 14.4 | 0.90 | 0.42 |
| Sexual Harassment Via E-communication | 69.7 | 86 | 16.4 | 13.9 | 65.9 | 75 | 20.0 | 14.1 | 0.15 | 0.92 |
| **Stalking Victimization** | 65.7 | 103 | 19.6 | 14.7 | 67.70 | 61 | 16.30 | 16.0 | 0.23 | 0.58 |
| **Dating Violence Victimization** | 65.0 | 78 | 14.9 | 20.2 | 68.5 | 37 | 9.9 | 21.6 | 0.029 | 0.42 |
| **Sexual violence victimization** |  |  |  |  |  |  |  |  |  |  |
| Sexual contact | 79.4 | 30 | 5.7 | 14.9 | 76.3 | 24 | 6.4 | 17.3 | 0.58 | 0.31 |
| Attempted sex | 83.4 | 10 | 1.9 | 14.7 | 77.3 | 12 | 3.2 | 19.5 | 0.16 | 0.06 |
| Attempted coercion | 84.2 | 7 | 1.3 | 14.5 | 78.9 | 11 | 2.9 | 18.1 | 0.07 | 0.14 |
| Attempted rape | 84.4 | 6 | 1.1 | 14.5 | 77.9 | 10 | 2.7 | 19.5 | 0.06 | 0.048 |
| Coercion | 81.5 | 20 | 3.8 | 14.7 | 66.4 | 12 | 3.2 | 30.4 | 0.93 | <0.001 |
| Rape | 81.5 | 12 | 2.3 | 16.2 | 65.30 | 11 | 2.9 | 31.7 | 0.27 | <0.001 |
| Any sexual violence victimization | 78.3 | 41 | 7.8 | 13.9 | 75.5 | 31 | 8.3 | 16.3 | 0.70 | 0.32 |
| **Sexual misconduct** | 26.3 | 294 | 56.0 | 17.7 | 25.6 | 196 | 52.3 | 22.1 | 0.79 | 0.09 |

The overall prevalence rate for sexual harassment victimization by students was 34.3%, with the sub-domain of sexual hostility being the most prevalent (24.1%), followed by sexist hostility (19.3%), sexual harassment via e-communications (18.6%), and unwanted sexual attention (18.3%) (Table 2). The overall prevalence rate of sexual harassment victimization by students did not differ by school or by gender (Tables 3 and 4).

Taken together, prevalence rates for sexual harassment based on reports by students were generally high. Compared to girls, boys more often reported experiencing sexual harassment victimization by faculty or staff. Compared to students at the other two schools, those from the low-ranked school more often reported unwanted sexual attention from faculty or staff (18.7%).

### Reports of stalking victimization

Almost 1 in 5 students (18.3%) reported experiencing stalking victimization since enrolling in school (Table 2). This prevalence rate did not differ significantly across the high-ranked school (21.8%), middle-ranked school (18.0%), and low-ranked school (15.2%) (Table 3). The prevalence of stalking victimization also did not differ significantly by gender (girls 19.6%, boys 16.3%) (Table 4).

### Reports of dating violence victimization

Forty-eight students (5.1%) reported not having had any boy or girlfriend since enrolling in school, and these students were excluded from this analysis. The prevalence rate of dating violence victimization among participants who reported to have had a boy or girlfriend was 13.1% (Table 2). This prevalence did not differ significantly across schools (Table 3). Girls more often reported dating violence victimization than did boys (14.9% vs. 9.9%) (Table 4).

### Reports of sexual violence victimization

The reported prevalence of unwanted sexual contact was 6.4%, and this prevalence did not differ across schools or gender. Survivors reported a prevalence of attempted sex at 2.8%, also similar across school and gender. The reported prevalence of sexual coercion was 4.1% (Table 2). This prevalence was highest in the middle-ranked school (6.2%), followed by the low-ranked school (4.4%) and the high-ranked school (1.6%), with the difference between the high-and middle-ranked schools being significant (Table 3). The prevalence of sexual coercion did not differ by gender (3.8% vs 3.2%) (Table 4). The reported prevalence of rape was 3.2%, which was similar across schools and between genders.

The prevalence of any sexual violence victimization experience was 8.7%, and prevalence rates did not differ significantly by school or by gender (Table 3 and Table 4).

### Overall sexual misconduct

The overall sexual misconduct was estimated if participants reported yes to any experience of victimization by sexual harassment, stalking, dating violence, and sexual violence. The overall prevalence was 54.5% (Table 2). High-ranked school reported the highest overall prevalence of sexual misconduct (60.6%), and the low-ranked school reported the lowest by 49.7% (p= 0.012) (Table 3). The prevalence rates of overall sexual misconduct did not differ by gender (Table 4).

## DISCUSSION

This pilot study is the first in Vietnam to collect comprehensive data on the forms of sexual misconduct experiences among students since their enrollment at three high schools in Ho Chi Minh city, Vietnam. We found a similar prevalence of sexual misconduct across three high schools of varying ranks. Between genders, boys experienced a significantly higher prevalence of sexual harassment by faculty or staff member, while girls experienced a significantly higher prevalence of dating violence victimization. To our knowledge, no previous study in Vietnam has explored a range of forms of sexual misconduct in high school students.

The overall prevalence of sexual misconduct was 54.5% in our study. Compared with another study that administered the ARC3 survey among 1,053 community college students in the U.S. (Howard et al., 2019), sexual harassment victimization by faculty or staff was higher in our study (40.2% vs 20.4%). In our study, sexist hostility was the most commonly reported form of sexual harassment victimization (30.2%). Compared to girls, boys more often reported unwanted sexual attention (18.7% vs. 10.9%, p 0.001) and sexual coercion committed by staff members (10.1% vs. 4.6%, p 0.001). While this gender difference was unexpected, it corroborates findings from two studies. First, a nationally representative school-based study of 10,410 Israeli students in Grades 7–11 showed boys more often experienced sexual maltreatment than girls (9.9% vs. 5.9%), including unwanted sexual behaviors and sexual coercion (Benbenishty et al., 2002). Second, a study of 1,376 Taiwanese students in grades 7-9 found that boys reported a higher rate of sexual victimization by teachers than girls (7.2% vs. 2.8%, p<0.001) (Chen & Wei, 2011). On the other hand, a study of 2,808 secondary school students in the Netherlands in 2005 found that girls experienced higher unwanted sexual behaviors by teachers than did boys (30.4% vs 20.3%, p<0.05) (Timmerman, 2005). The lower rate of sexual harassment by teachers among girls than boys could be due to girls’ limited knowledge of consent and the range of behaviors that constitute sexual misconduct, their hesitancy to report sexual misconduct out of shame or fear of blame, or their unwillingness to challenge norms of male sexual privilege and gendered expectations that girls should maintain social harmony (Ali et al., 2021; Bergenfeld, Tamler, et al., 2022; Lewis et al., 2022)

Sexual harassment victimization *by students* in our study also was higher (34.3%) than in the study by (Howard et al., 2019) (27.9%) but lower than in a 2011 study of 1,965 students in grades 7-12 in the US (48%) (Hill & Kearl, 2011). Notably, estimated rates of sexual harassment found in our study fall within the ranges identified in a global systematic review of studies conducted in educational settings (14.4%-73%), some of which relied on similar scales to measure sexual harassment (Ranganathan et al., 2021).

The level of stalking victimization in our study (18.3%) was only slightly higher than among community college students in the U.S. (14.3%) (Howard et al., 2019) and similar to estimates among 725 students attending UK universities in 2020 (16%) (Bull & Bradley, 2023). In addition, ours and the UK study found that girls experienced a higher prevalence of stalking than did boys, though the difference was not statistically significant in our study.

The prevalence of dating violence was lower in our study (13.1%) than among community college students in the US (17.9%) (Howard et al., 2019) and university students in the UK (26%) (Bull & Bradley, 2023). The higher prevalence of dating violence among girls than boys in our study corroborates findings in the UK study of university students (Bull & Bradley, 2023), the Youth Risk Behavior Survey in the US (Clayton et al., 2023), and a study of physical and psychological dating violence in Quebec (Hébert et al., 2017).

Sexual violence in our study (8.7%) was reported less often than among US community college students (11%) (Howard et al., 2019), less often than among participants in the Youth Risk Behavior Survey in the US (11%) (Clayton et al., 2023), but more often than among 2,200 college students in China (4.2%) (Wang et al., 2015).

Lower rates of dating violence and sexual violence in our study than in other studies could be due to age differences in the samples, and correspondingly, the lower prevalence of dating among high-school students in Vietnam than among university students in Western settings. Although expectations for sex and sexual activity are increasing among young people in Vietnam (Bergenfeld, Tamler, et al., 2022), students still may be less sexually active, and thus, would less often be exposed to sexual violence in casual or dating relationships (Ansari-Thomas et al., 2020; Li et al., 2010).

Our study has some notable strengths. First, this study is the first to explore a range of forms of sexual misconduct among high school students in Vietnam. Thus, the high prevalence of sexual misconduct victimization in this population fills an important knowledge gap. Second, the ARC3 survey includes numerous questions to identify each form of sexual misconduct, and item sets capturing each form exhibit good psychometric properties in US university populations (Swartout et al., 2019),(Tilley et al.,

2020). Third, our sample size of 754 students is large enough to detect some statistically significant differences across high schools and genders, respectively.

Some limitations of the study offer opportunities for future research. First, data on sexual misconduct did contain some missingness, for which several explanations are plausible. First, some non-participation may have been due to collecting data in May, overlapping with the examination period. Second, higher rates of missingness were observed for later questions in the survey, which also focused on sexual violence victimization (Table 2). This higher level of missingness may have been due in part to respondent burden, as suggested by the higher levels of missingness in later modules (e.g., 16.2%-22.7% for sexual violence victimization) than in earlier modules (e.g., 10.5% to 13.8% for sexual harassment). Higher missingness also may have been due to the sensitive nature of some questions, such as ones about rape, for which 22.7% of responses were missing. Third, some duplicate records were created due to challenges with internet connectivity, which may have contributed to the missingness in later modules because participants with connectivity issues has to re-enter REDCap through a new ID and re-complete the survey. Despite this limitation, rates of missingness in our study are similar to those found in a campus climate survey of undergraduate students across nine universities (Krebs et al., 2016).

## CONCLUSION

The prevalence of sexual misconduct victimization is high among 11^th^ and 12^th^ grade students attending three high schools in Ho Chi Minh City. Our estimates provide an evidence-base for dialogue with parents, school administrators, and youth to understand the scale of the problem and need for preventive and response strategies in high schools. The ARC3 survey warrants psychometric and invariance testing across students in Vietnam to ensure that the comparisons made here are valid. In that case, findings provide a critical evidence base to develop educational programs that support the primary prevention of sexual misconduct in high schools across Ho Chi Minh City and nationally.

## Data Availability

All data produced in the present work are contained in the manuscript

## Acknowledgments

The authors thank Ms. Quach Thu Trang for reviewing the ARC3 questionnaire in Vietnamese.

## Author contributions

Conceptualization: KTT, KMY. Methodology: KTT, RML, KMS, MHT, OT, KMY. Formal analysis: KTT. Investigation: KTT. Resources: KTT, KMS. Data Curation: KTT. Writing - Original Draft: KTT, KMY. Writing - Review & Editing: KTT, RML, KMS, MHT, OT, KMY. Visualization: KTT, KMY. Supervision: KMY. Funding acquisition: KMY. All authors read and approved the final manuscript.

